# Effect of laser assisted local anesthesia in single-visit root canal treatment for mandibular molar teeth with acute irreversible pulpitis

**DOI:** 10.1101/2021.08.03.21261519

**Authors:** V Subashri, V Nivedha, Anand Sherwood, Paul V Abbott, James L Gutmann, Cert Endo, Omar Farooq, MV Aarthi

## Abstract

Present study evaluated the efficacy of laser activation to control intra- and post-operative pain in single-visit root treatment for mandibular molar teeth with acute irreversible pulpitis following 2% lignocaine inferior alveolar nerve block. Ninety-eight patients presenting with pain were randomly divided into two anesthetic groups. Group-I inferior alveolar nerve block plus buccal infiltration and intra-ligamentary injections, Group-II inferior alveolar nerve block followed by laser irradiation focused directly on the pulp tissue. Intra- and post-operative pain intensities were assessed on a 10-point scale.The mean intra-operative pain scores in group-I was 6.62 ± 1.6 and in group-II before and after laser irradiation pain scores was 6.94 ± 2.1 and 1.3 ± 2.04, respectively. Post-operative pain scores at 24-hrs in the laser group were significantly higher. Laser irradiation applied directly on pulp tissue for control of intra-operative pain was effective, thereby negating the need for additional local anesthesia.

**Clinical relevance:** Laser activation was effective method to control intra-operative pain in irreversibly inflamed pulp.

Laser irradiation did not cause adverse post-operative pain.

## Introduction

A mandibular molar tooth with acute irreversible pulpitis is one of the most disconcerting situations to be encountered in an endodontic clinic (1). Treatment options for these situations are limited to either pulpotomy, pulpectomy, extraction or prescribing potent analgesics (1). Achieving satisfactory anesthesia and reducing the incidence of post-treatment discomfort are the particular difficulties encountered in the endodontic treatment of these teeth (1). Multiple strategies have been exploredto attain profound anesthesia and to control post-operative pain in these situations (2). An earlier report from the authors’ department concluded that pre-operative ketorolac tromethamine was not effective in reducing the intra-operative pain for mandibular molar teeth with acute irreversible pulpitis and apical periodontitis when using inferior alveolar nerve blocks (IANB) with both lignocaine and articaine anesthetic agents. However, it was effective in reducing post-operative pain after using lignocaine which was in agreement with a meta-analysis (3, 4). Mean intra-operative pain scores for both anesthetic agents were 4.33 and 4.22, respectively, and 27 (21.4%) patients required supplemental anaesthesia to control their intra-operative pain in the aforementioned study (3). In another clinical study in the authors’ department where an inferior alveolar nerve block was used plus buccal nerve infiltration and intra-ligamentary injection with both anesthetic agents for single-visit root canal treatment in mandibular molar teeth with acute irreversible pulpitis, the mean intra-operative pain scores were 2.43 and 3.19 for the two anesthetic agents, respectively, and 12 (9.4%) patients required supplemental anesthesia (5).Supplemental anaesthesia requirements were far more reduced in the above-cited clinical trial when patients were given multiple injections prior to commencement of the root canal treatment (5).

Patients are generally apprehensive regarding multiple injections before root canal treatment. Furthermore, the patients’ medical condition may preclude additional local anaesthetic agents. To circumvent this problem, the use of laser activation was investigated in a pilot study. Laser application for intra-operative pain control in a previous clinical trial was used on the hard tissue of the tooth prior to local anesthetic deliveryand never directly on the pulp (6, 7). However, laser application is known to allow painless soft tissue excision. Laser activation directly on the pulp to reduce intra-operative pain in acute irreversible pulpitis after local anesthetic administration has not yet been investigated.

The aim of the present study was to assess laser application efficacy when applied directly on the pulp in patients with intra-operative pain following inferior alveolar nerve block in mandibular molar teeth with acute irreversible pulpitis. The secondary aim was to assess their post-operative pain following laser application in single-visit root canal treatment.

## Materials and Methods

A total of 172 patients requiring root canal treatment for pain due to acute irreversible pulpitis with primary acute apical periodontitis in carious mandibular first and second molars were identified based on standard subjective and objective criteria (8).The study was conducted from February 2020 to January 2021. The sample size was determined from the assumption of a pilot study conducted in the corresponding author’s department in 20 patients managed with a similar protocol where there was intra-operative pain reduction of 80%. Using a χ^2^with effect size = 0.50 and power (1 - β err prob) = 0.95 (G*Power 3.1.9.2, Universität Kiel, Germany), 80 patients were required. Accounting for attrition in the follow-up, the number was increased to 98 patients. Subsequent to approval from the Institutional Ethics Committee, the trial was registered with ClinicalTrials.gov and a total of 172 patients with acute irreversible pulpitis and primary acute apical periodontitis due to carious mandibular molar teeth were enrolled in the study. The patients (or where appropriate, parents or guardian) were informed about the nature of the treatment and the study, and they were asked to sign an informed consent form. The methodology adopted for this study was similar to clinical experiments conducted earlier in the authors’ department (3).

Patients referred to the Department of Conservative Dentistry and Endodontics with pain due to acute irreversible pulpitis from carious mandibular first and second molar teeth requiring root canal treatment were evaluated as possible candidates for this study. Subjects aged between 13 and 70 years were included in the study. All patients reported mild to severe pain that was continuous, spontaneous, radiating, nocturnal or throbbing in nature. All the teeth included in this study responded to cold pulp sensibility testing (Endo-Frost, Coltene Whaledent, Switzerland) with exaggerated pain, with or without lingering. They also had tenderness on percussion. Additionally, profuse bleeding was evident upon gaining access into the pulp chamber. The teeth included in the study also did not have any evidence of periapical bone changes on the pre-operative periapical radiographs.

Exclusion criteria were absence of pain upon gaining access to the pulp following local anesthetic administration, teeth with poor periodontal or restorative prognosis, patients with systemic ailments or conditions hindering single visit root canal treatment, patients not willing to participate in the post-operative recall evaluation, any anatomic variation such as extra roots or root canals, C-shaped roots, and patients with a history of allergy. A PRIRATE 2020 (9) flow chart of the experimental procedure is depicted in Figure 1.

**Figure 1.**
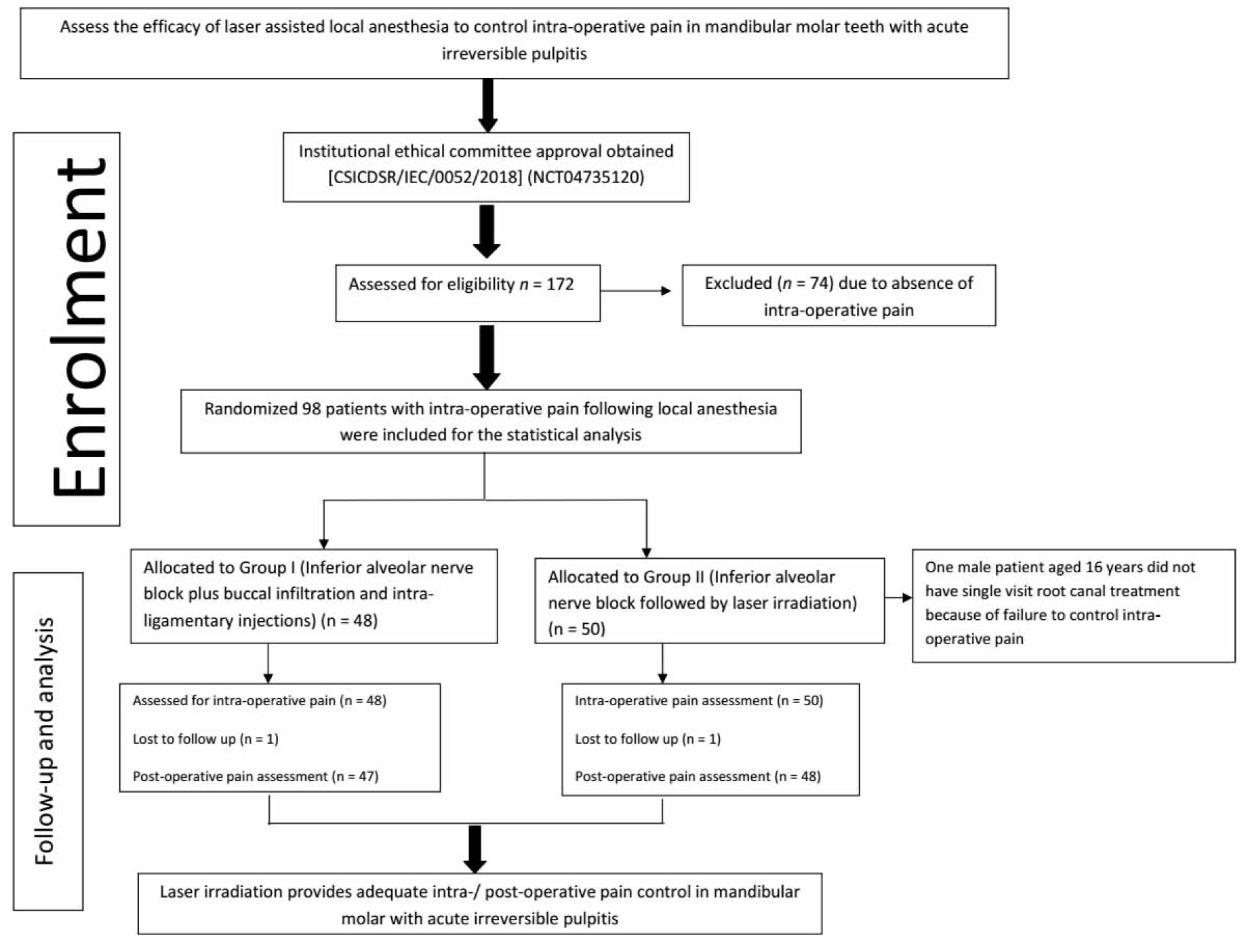
PRIRATE 2020 (9) flowchart illustrating the flow of participants.

### Experimental procedure

All root canal procedures were done by a single operator. The levels of pre-, intra- and post– operative pain at 24 hrs and 48 hrs for each patient were recorded using a 10-point visual analogue scale (VAS). The participants indicated the intensity of their pain by choosing a number using the following values: levels 1–3, mild pain; levels 4–7, moderate pain; and levels 8–10, severe pain. Patients were assisted in their pain assessment where necessary by an independent and calibrated endodontist.

The patients were randomly allotted to two local anesthetic groups, Group I – Inferior alveolar nerve block (IANB) plus buccal infiltration and intra-ligamentary injection, and Group II – Only IANB followed by laser irradiation. Randomization was done by an independent endodontist not associated with treatment delivery who picked concealed lots written with group names.

### Group I

An intra-dermal injection of 0.2 mL of the local anesthetic agent to be used was given prior to the IANB in order to rule out any allergy to the anaesthetic agents. Then, 2.5 mL of 2% lignocaine containing 1:80,000 adrenaline (Lignox, Warren Pharmaceuticals, Mumbai, India) was administered as an inferior nerve alveolar nerve block (IANB) plus a 1.5 mL for a buccal infiltration and 0.1 to 0.2 mL as an intra-ligamentary injection using the same anesthetic agent. The intra-ligamentary injections were given at four sites for each tooth on the buccal and lingual sides. The local anesthesia was administered seven minutes prior to commencing the root canal treatment procedure. Cold pulp sensibility tests and percussion evaluations were performed after inquiring about the level of lower lip numbness and before the access cavity opening were commenced. Responses to these tests were recorded. If sufficient anesthesia was not attained, an additional IANB was administered with the same local anesthetic agent used previously.

Intra-operative pain was recorded at only one instance with instructions that if pain was felt at any stage of the root canal treatment procedure, the patients were asked to raise their left hand. The intensity of this intra-operative pain was recorded, and patients were re-assessed regarding the need for either supplementary intra-ligamentary or intra-pulp anesthesia, or an additional IANB. If supplementary local anesthetic was required, the same local anaesthetic agent used for the original IANB was employed.

### Group II

Laser application for control of intra-operative pain is a novel technique with no previous investigations to refer to so the pulse rate was assessed as an objective evaluation of the pain control in group II only. Prior to local anesthesia, each patient’s basal pulse rate (PR1) was determined using a finger pulse oximeter (Oxywatch, Beijing Choice Electronic Technology Co, Ltd, Beijing, P.R. China). An intra-dermal injection of 0.2 mL of local anesthetic agent was given prior to the inferior alveolar nerve block (IANB) to rule out any allergy to the anesthetic agent. IANB local anesthesia with 2.5 mL of 2% lignocaine containing 1:80,000 epinephrine (Lignox, Warren Pharmaceuticals Pvt Ltd, Mumbai India) was administered seven minutes prior to commencing the root canal treatment procedure. Cold pulp sensibility tests and percussion evaluations were done as in previous group prior to access cavity preparation. Upon access to the pulp,the intensity of intra-operative pain (IP 1) using the VAS score and the pulse rate (PR 2) were determined. Following this, an InGAsP semiconductor diode-based laser (iLase, Biolase, CA, United States) was applied to the pulpboth in the pulp chamber and inside the root canals. Activation was with 940 nm wavelength, 1.5 Watts for 60 to 180 seconds in continuous mode for each canal orifice with an E2–20 tip (200 µm diameter and 20 mm length tip). Following laser activation, root canal treatment was commenced with instructions for the patients to raise their left hand if they felt pain – if they did feel unbearable pain, then further intra-operative pain (IP 2) and pulse rate (PR 3) values were recorded. The patients were re-assessed about the need for either supplementary intra-ligamentary or intra-pulp anesthesia, or an additional IANB. If supplemental or additional local anaesthetic was required, the same local anaesthetic agent used for the original IANB was used. Patients experiencing no pain after laser activation were assessed for IP 2 after completion of root canal instrumentation and their pulse rate (PR 3) was also recorded at that stage. Intra-operative pain assessments in both groups were done by an independent, calibrated endodontist.

### Root canal treatment procedure

Working length was determined using aRoot ZX Mini Apex Locator (J Morita, Kyoto, Japan) and Flexer files (Mothers Dental International, Adelaide, Australia) were used for root canal preparation according to the manufacturer’s instructions using an EndomateTC2 motor (NSK Inc., Tochigi, Japan). Canal lubrication and smear layer management were done with EDTA (10%) and carbamide peroxide (15%) (Endoprep RC, Anabond Stedman Pharmaceuticals, Chennai, India). Sodium hypochlorite (3%) (Septodont Healthcare India Pvt. Ltd, Raigad, India) was used duringroot canal preparation. In all teeth, the sodium hypochlorite irrigation with upto 5 mL for each canal was used. Initial irrigation of the root canal was done after establishing a glide path upto size 20 or 25K-file (Mani, Co., Tokyo, Japan). Mid-rinse and final irrigation followed the use of the rotary instruments. A total of 7 mL of irrigating solution was used in each canal during the treatment. The irrigation solution was delivered into the root canals using a side-vented 25-gauge needle (RC Twents, Prime Dental Products, Mumbai, India) with a standard syringe. The needle was inserted as far apically into the canal as possible but without any binding within the canal. Gentle force was used on the syringe to deliver the irrigant, and the needle was moved up and down inside the canal to assist with irrigant flow and to ensure no binding of the needle to the canal walls.

Upon completion of the root canal preparation to an apical size of either 6% size 20 or 25, apical patency was checked using a size 10 K-file (ManiCo., Tokyo, Japan). Root canal filling was done using a greater taper single gutta-percha cone (DiaDentGroup, Seoul, Korea), along with a zinc oxide eugenol-basedcement (Prime Dental Products, Thane, India). A post-operative radiograph was taken to ensure the canals were filled tothe working length and there was no extrusion of filling material into the periapical tissues. All the patients were prescribed paracetamol 500 mg as analgesics but the patients were advised to only take them in the event of significant pain. The occlusion was not relieved in this study.

The post-operative pain levels and the need for analgesics were recorded after 24 and 48 hrs by telephoning each patient.The post-operative pain enquiry was made by the same independent endodontist who inquired about intra-operative pain. If analgesics were required, the patients were questioned about which medication they used, the dosage, how often they had taken them and whether they were effective. Furthermore, the trigger mechanism of any post-operative pain in group-II was recorded as spontaneous continuous pain, spontaneous occasional pain, or stimulus-based pain. Trigger mechanism was evaluated for group-II only as the laser usage was attempted for first time and how it matches with other previous investigations.

### Statistical analysis

Statistical analysis was performed using IBM SPSS software version 23 (IBM Corp., Washington, USA). Normality of pre-, intra-(IP – 1 and IP – 2) and post-operative pain scores were checked by theShapiro–Wilk test. The data were skewed and deviated from normal distribution - therefore, the comparisons of these values were done by non-parametric Wilcoxon signed rank, Mann-Whitney and Kruskal–Wallis tests. Pulse rate increase from PR1 to PR2 and pain experience from IP1 to IP2 were calculated as percentages for group-II, as follows:

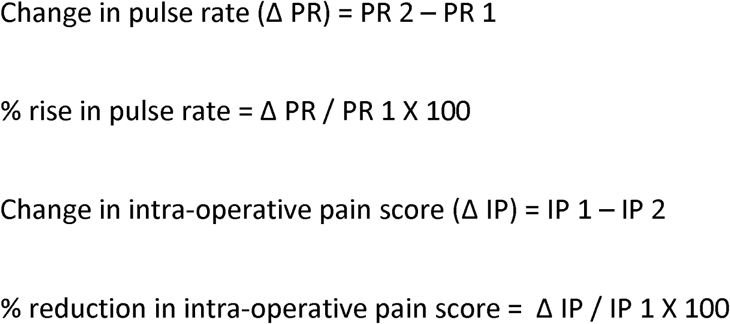

The level of significance was setat 5%.

## Results

A total of 172 patients enrolled in the study and of these, 98 patients with intra-operative pain following local anesthesia (42 males and 56 females, aged between 13 to 70 years) were included for the statistical analysis. In group I and II, 48 and 50 patients were allotted respectively. The overall mean pre-operative pain score was 6.92 ± 1.78 with no significant difference between the two genders (Mann-Whitney test). Cold test responses after local anesthetic administration did not show a significant association with intra-operative pain (Kruskal-Wallis test). In group II, one male patient aged 16 years requiring root canal treatment for his mandibular first molar did not have the treatment completed in the first appointment because of failure to control his intra-operative pain with laser application and supplemental local anesthesia.

### Group I

The mean pre-operative score was 6.62 ± 1.6. The mean intra-operative pain score was 3.25 ± 2.20. No significant difference was observed in intra-operative pain among the two genders (Mann-Whitney test). Three patients (2 males and 1 female) (6.3%) required supplemental injections for control of intra-operative pain. Mean intra-operative pain score for patients requiring supplemental anesthesia was 7.66 ± 2.51 and was significantly different (*P*< 0.05) according to the Mann-Whitney test compared to patients not requiring supplemental injections. None of the patients required additional IANB. One patient did not respond for post-operative evaluation until the completion of the trial. The mean 24 and 48 hrs pain scores were 0.89 and 0.31, respectively, with no significant gender difference.Thirteen patients (27.1%) required post-operative analgesics.

### Group II

The mean pre-operative pain score was 7.2 ± 1.91. The mean IP 1 score was 6.94 ± 2.1, PR 1 and PR 2 was 82.2 ± 14.3, 86.3 ± 15.9 respectively with no significant differences for either genders (Mann-Whitney test).The mean intra-operative pain score after laser application (IP 2) was 1.3 ± 2.0 and PR 3 was 80.1 ± 12.7 with no significant difference between thetwo genders (Mann-Whitney test). The Wilcoxon-Signed rank test showed a significant difference (*P = 0*.*00*) between the intra-operative pain scores before and after laser activation. The basal pulse rate (PR 1) was significantly different (*P = 0*.*00*1) to the rate upon access opening (PR 2) according to the Wilcoxon-Signed rank test. This rise in percentage was 4.96%. Following laser activation, the percentage reduction of intra-operative pain was 81.3% which was significantly different (*P = 0*.*00*) (Wilcoxon-Signed rank test) (Fig.2). The reduction in pulse rate (PR 3) after laser activation from PR 2 was significant (*P = 0*.*00*) (Wilcoxon-Singed rank test).The pulse rate rise from the basal pulse rate did not have a significant association with intra-operative pain prior to laser application (Kruskal-Wallis test).

Four patients (2 males and 2 females) (8%) required supplemental local anesthesia; this was not significantly different from group-I (Mann-Whitney test). All supplemental injections were given intra-pulpally. None of the patients in this group also required an additional IANB. The mean IP 1 and IP 2 scores for the patients requiring supplemental anesthesia were 10 ± 0.0 and 6.50 ± 4.12 and these were significantly different from the scores for patients not requiring supplemental injections (*P*< 0.05) in Mann-Whitney test (Table 1).

**Table 1.** Intra-operative scores before and after laser application in patients requiring and not requiring supplemental injectionin group II.

| | | Mean intra-operative pain prior to laser activation ± Standard deviation | Mean intra-operative pain experience after laser activation ± Standard deviation | Number of patients |
| --- | --- | --- | --- | --- |
| Supplemental local anesthetic requirement | Yes | 10.00 ± 0.00 | 6.50 ± 4.12 | 4 |
| | No | 6.67 ± 1.97 | .8478 | 46 |
| Total | | 6.94 ± 2.10 | 1.30 ± 2.04 | 50 |

The post-operative pain incidence in this group was 47.9 % (23 patients) with no significant gender difference (Table 2). The post-operative pain incidence had no significant association with pre-operative pain type or the requirement for supplemental anesthesia similar to the earlier group. The mean post-operative pain scores for group-II after 24 and 48 hrs were 1.95 ± 2.4 and 0.8 ± 1.56, respectively. One patient in group-II did not respond and return for post-treatment assessment until completion of this study. Sixteen patients (33.3 %) required post-operative analgesics, and most of these patients required only one dose of analgesic to control the discomfort as in group-I. The trigger mechanism graph (Fig 3) shows the spontaneity nature of the post-operative pain (19 patients (82.61 %)) which made patients report post-treatment discomfort.

**Table 2.** Post-operative pain incidence among the two genders in group II

|  | Post-operative pain |  | Total |
| --- | --- | --- | --- |
|  | Yes | No |  |
| Male | 9 (42.9%) | 12 (57.1%) | 21 |
| Female | 14 (51.9%) | 13 (48.1%) | 27 |
| Total | 23 (47.9%) | 25 (52.1%) | 48 |

**Figure 2.**
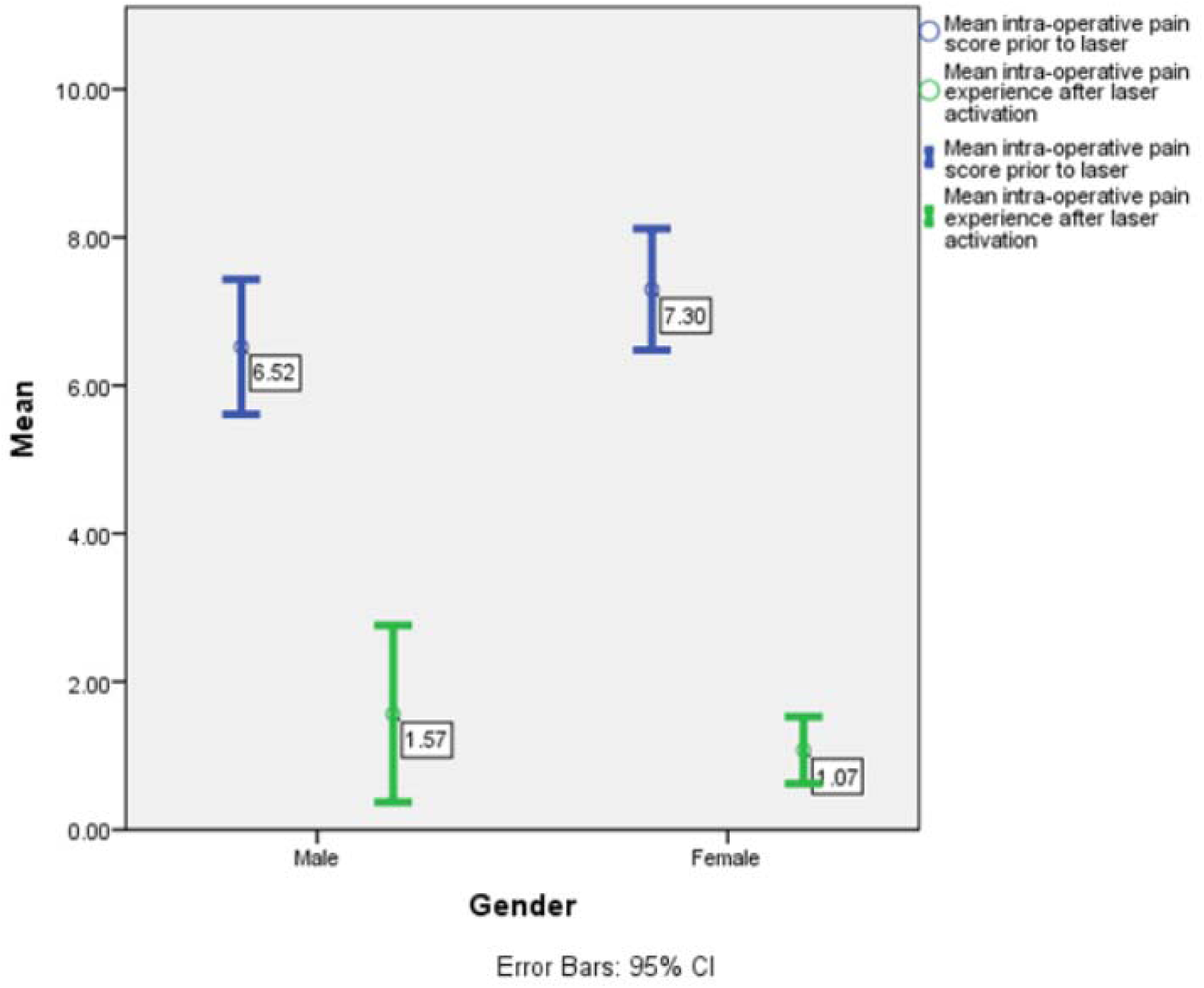
Mean intra-operative pain scores with error plots prior to and after laser irradiation in group-II.

**Figure 3.**
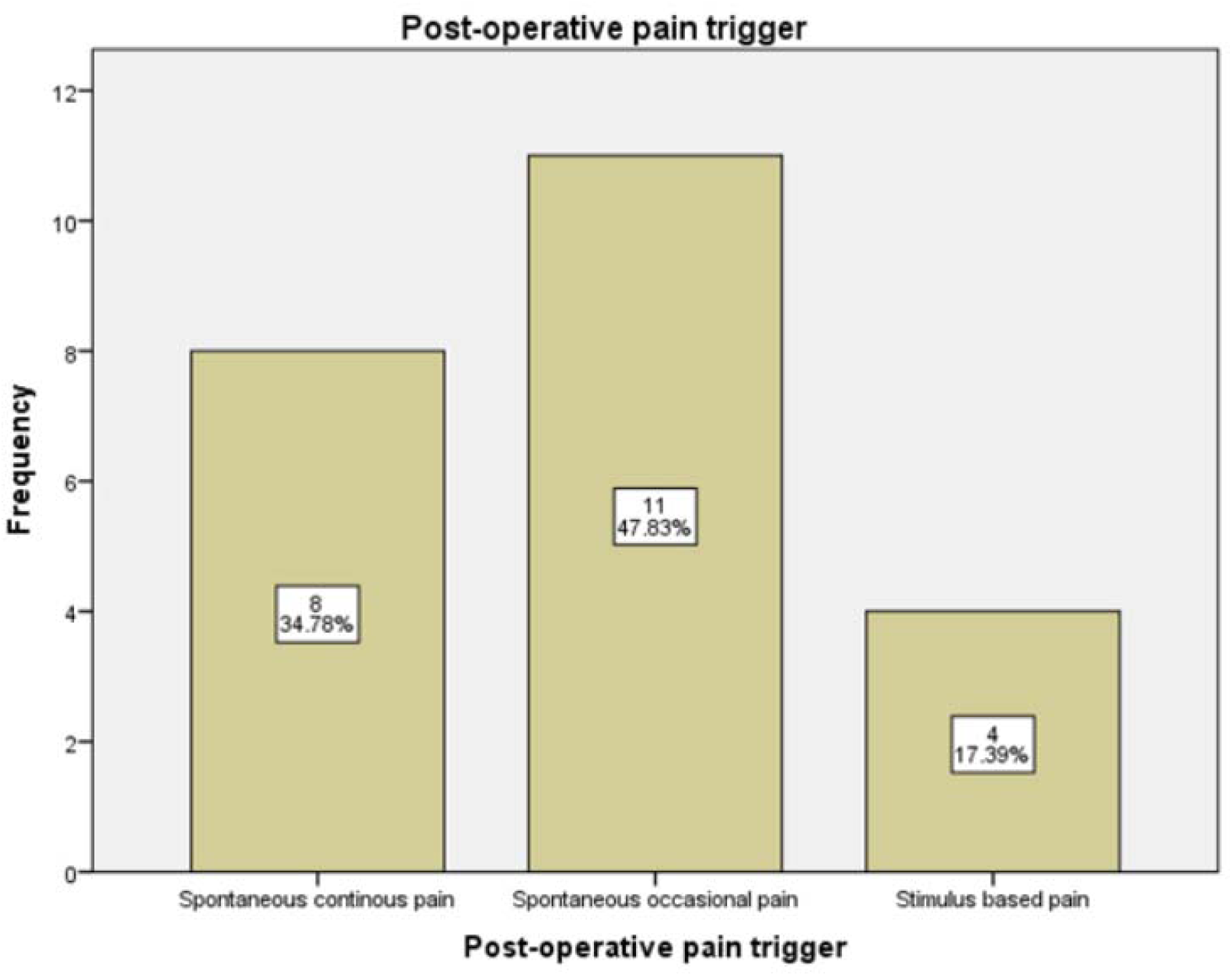
Post-operative pain triggers in patients with post-treatment discomfort in group-II.

### Inter-group comparison

The mean pre-operative pain score was similar for both groups (Mann-Whitney test). Mean intra-operative pain score after laser application (IP 2) was significantly (*P*< 0.05) less compared to group-I intra-operative pain score (Mann-Whitney test) (Table 3). Post-operative pain at 24-hrs scores for group-II patients were significantly (*P*< 0.05) higher compared to group-I (Mann-Whitney test)(Fig 4).

**Table 3.**
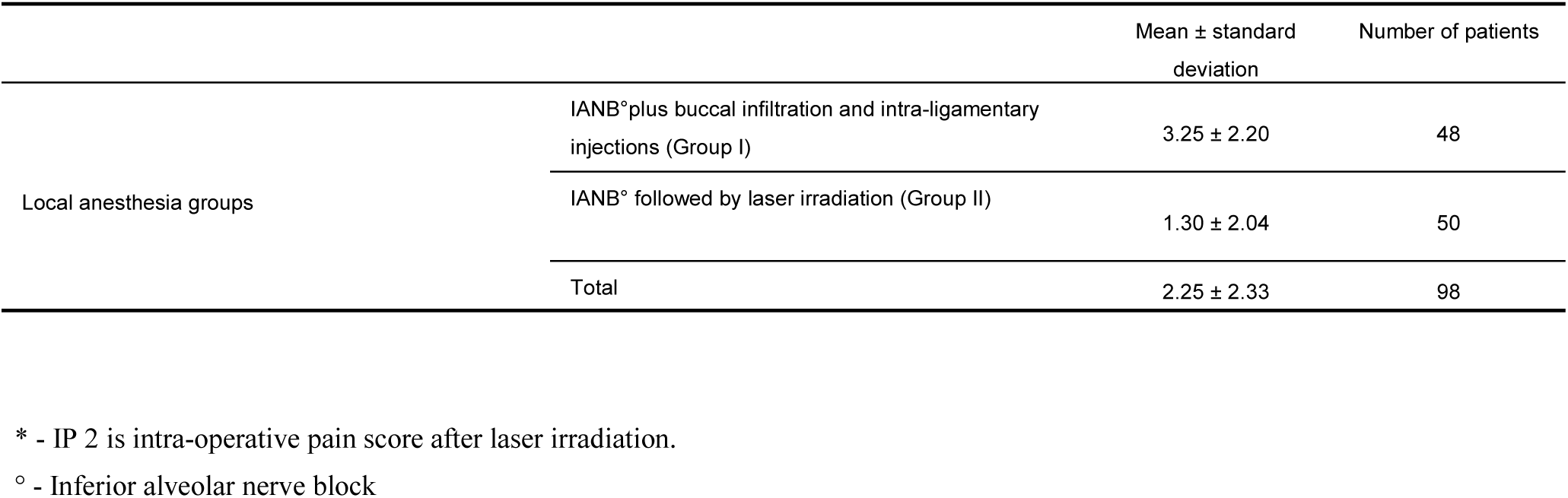
Mean intra-operative pain scores in group I and group II (IP 2*).

| | | Mean ± standard deviation | Number of patients |
| --- | --- | --- | --- |
| Local anesthesia groups | IANB<sup>°</sup>plus buccal infiltration and intra-ligamentary injections (Group I) | 3.25 ± 2.20 | 48 |
| | IANB<sup>°</sup> followed by laser irradiation (Group II) | 1.30 ± 2.04 | 50 |
| | Total | 2.25 ± 2.33 | 98 |

\* - IP 2 is intra-operative pain score after laser irradiation.
<sup>°</sup> - Inferior alveolar nerve block

**Figure 4.**
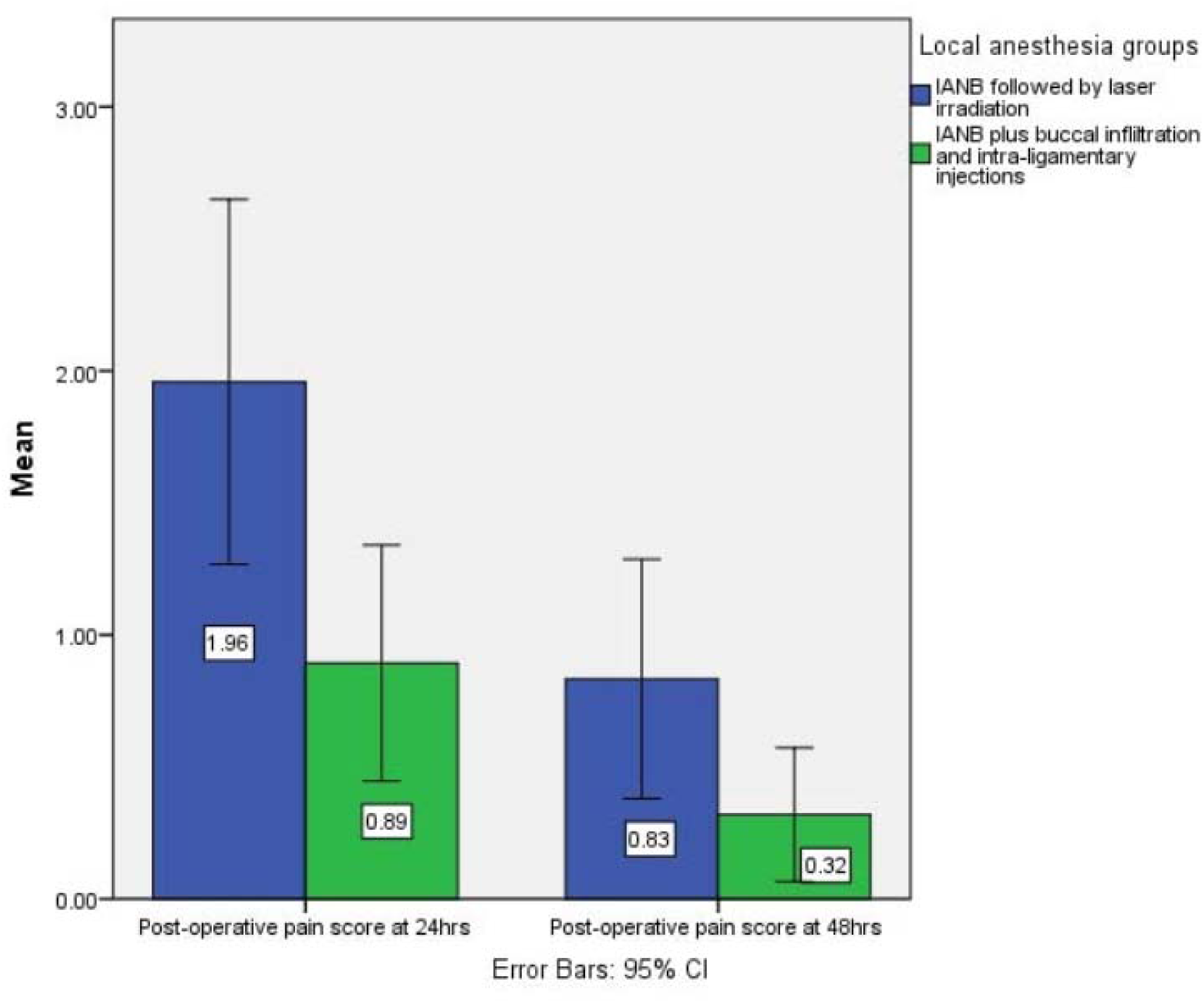
Post-operative pain score at 24 and 48 hrs for group I and II with error plots.

## Discussion

The primary aim of this study was to evaluate the efficacy of laser activation in reducing the intra-operative pain intensity and thereby indirectly reducing the need for supplemental anaesthesia. Only mandibular molar teeth exhibiting acute irreversible pulpitis were considered, as these teeth are the most difficult teeth to anesthetize with a single IANB injection (3). The result of the present study highlights the efficacy of a laser to reduce both the intra-operative pain intensity and the need for supplemental anesthesia. A previous study from the authors’ department (3), using a similar study design and a single 2% lignocaine IANB injection indicated that the mean intra-operative pain score for patients with intra-operative pain was 4.6 ± 2.4. Furthermore, the requirement for supplemental anesthesia was 23.8 % (15 patients), which was far greater than the group-II of the present study (IP2 - 1.3 ± 2.0; supplemental injection requirement - 4 (8%) patients). Additional IANB was required in 9 (14.3 %) patients in the above cited study (3) compared to none of the patients requiring this intervention in the present study.The mean intra-operative pain score (IP 1) in group-II prior to laser application (6.94 ± 2.1) is higher than the earlier mentioned study and this may be due to the pre-operative oral ketorolac tromethamine administration in that study (3). Present data supports the efficacy of a laser in controlling even the elevated intra-operative pain intensity without pharmacological aid despite the higher pre- and intra-operative pain intensity.

In this study, patients administered with additional buccal nerve infiltration and intra-ligamentary injections coupled with the IANB injection had a mean intra-operative pain score of 3.25 ± 2.20 which was significantly higher than the laser group (IP 2) despite the elevated pre-operative pain levels observed in the laser group. Supplemental injections were required in 4 patients (8%) in the laser group-II which was similar to group-I. These numbers indicate the efficacy of laser activation in controlling the intra-operative pain without the need for additional or supplemental injections.

A 10-point VAS was employed as our previous studies with irreversible pulpitis (3, 5, 10, 11) followed this similar protocol, which allowed us to have standardised metrics for efficient patient communication. Additionally, most of the patients encountered in the authors’ department have limited education which would greatly impair their interpretation of a pain scale with more than 10-points. Telephonic recording of post-operative pain assessment was adopted as post-treatment compliance of patients for recall visits is very poor once the pre-operative symptoms subside.

Pulse rate and intra-operative pain changes were measured to provide an accurate and informed metric tool relative to the expected change in these values, while performing the treatment.The pulse rate increase from the basal level (IP 1) to just before laser application (IP 2) was 4.96 % and this was independent of the intra-operative pain intensity. This implies that the pulse rate change was not a reliable tool for evaluating intra-operative pain experience. Reduction percentage of intra-operative pain intensity after laser activation was 81.26 %,which provides a valuable source of information in patient management.

Laser intensity of 1.5 watts in continuous wave mode for 60 to 180 secs was adopted in the present study based on previous research (6, 7). In that research 1.5 watts of laser energy for 1 min produced a reversible intra-dental nerve conduction block without contacting the pulp tissue (6). They also indicated that total laser energy induced analgesia acts via suppression of intra-dental nerve responses to mechanical and electrical stimulation (6).

The present study varies from that of Chan *et al. (6)* in that the laser energy here was used to anesthetize irreversibly inflamed pulps and also with direct contact on the pulp tissue that required an extended length of time of laser application. A wide range in time of laser application was adopted in this study due to the lack of previous scientific evidence to know the precise time that would be required. Diode laser application has been long associated with painless treatment in minimally invasive soft tissue surgical procedures with minimum collateral damage to either the soft tissue or the bone in different modes of direct contact (12).The direct contact mode of diode lasers on soft tissues has been shown to produce tissue vaporization and thermal necrosis.Therefore,the reduction in intra-operative pain observed in the present study could not be linked to suppression of intra-dental nerve conduction block (12).

Intra-pulp anesthetic injections achieve their anesthetic effect due to the physical mechanism of back pressure, irrespective of the type of anesthetic agent, rather than by pharmacological means (13). Laser assisted intra-pulp anesthesia in the present study could also be due to the physical effects of tissue vaporization and thermal necrosis instead of an alteration of any specific neurological conduction mechanism. No direct comparison of the present results with other studies could be made as a literature search failed to identify any similar attempts to assess lasers for irreversible pulpitis. Earlier studies on laser application for the management of irreversible pulpitis to control intra- and post-operative pain have been done using laser irradiation on the external tooth surface, either before or after root canal treatment, but not directly on the pulp during the treatment procedure (7, 14, 15).

Supplemental anesthesia was required for 4 patients in laser group who had a mean intra-operative pain score of 10 (severe) prior to laser activation and 6.5 (moderate) after laser activation where the reduction achieved was not close to the reduction achieved for other patients. One reason for this occurrence was that the length of the laser fiber tip was only 20 mm, which needed to be bent to enter the canal orifices. The cases requiring supplemental anesthesia had root canal lengths of more than 22 mm which prevented the fiber tip from directly contacting the entire pulpto achieve the anesthetic effect as explained previously. In the future, laser fiber tip manufacturers should customize their tips with different lengths for use in anesthetizing the pulp within the root canals.This further supports the earlier explanation of a physical mechanism behind the laser assisted pulp anesthesia.

In this investigation, intra- and post-operative pain was assessed by an independent endodontist whereas the laser activation and root canal treatment was done by the same operator. This was the case as there were no precursor studies to refer to in order to know the intensity, time of duration or mode of laser irradiation for achieving the pulp anesthesia. Hence, it was preferred that the same clinician operate the laser to allow for real time monitoring of patient discomfort during root canal treatment and altering of the above mentioned parameters as and when required for each case. The present study also did not allow for a direct placebo or sham laser application since the patients had acute irreversible pulpitis and an earlier investigation (3) with similar conditions showed a single 2.5 ml of lignocaine IANB was not sufficient to achieve complete pulp anesthesia to allow completion of the root canal treatment. As a consequence, a placebo or sham laser group would have meant subjecting the patients to unnecessary discomfort and also might have led to abandoning the root canal treatment in the first appointment. This statement can be appreciated by observing that the mean intra-(IP 1) and pre-operative pain values in group-II were almost identical.

The secondary objective of the study was to investigate post-operative pain following laser application to help understand the adverse effects posed by this technique. Post-operative pain scores at 24-hrs for laser group were significantly higher compared to group-I. The post-operative pain incidence in the laser group was 47.9%, which was higher than the earlier study (3) of 16.7%. However, as discussed above, this earlier study had pre-operative oral analgesic administration to control post-treatment discomfort. However; the 24-and 48-hrs post-operative pain intensity was similar to the earlier study (3). Another study (11) from the authors’ team that focused on anesthetizing teeth with acute irreversible pulpitis and research published by Arias *et al*.*2013 (*16) without pre-treatment analgesic administration, have shown comparable post-operative pain incidences to the present study. Likewise, the

post-treatment pain intensity was also similar. Laser irradiation inside the root canal system has been associated with minimal potential to cause damage to periodontal tissues or bone (17). This implies that the post-treatment data with laser application did not cause any adverse post-treatment effects when used to attain pulp anesthesia despite the significantly higher post-operative pain intensity associated with the laser group. Arias *et al. 2013 (*16) reported that spontaneously triggered post-operative pain was the most commonly reported type of discomfort, which is in agreement with the current results. Occlusal contact was not relieved in the present study inorder to stimulate the worst-case scenario for evaluating post-operative pain as the presence of occlusal contact has been associated with increased post-treatment discomfort (10, 16). Despite this, the post-operative pain occurrence was similar to other reports on pain following root canal treatment (16), thereby confirming the efficacy of laser application.

In conclusion, laser activation for the control of intra-operative pain reduced the requirement for supplemental pharmacological support. Further investigations in the authors’ department with a larger sample size for evaluating the efficacy of pre-operative oral analgesic intake to control the post-operative pain in laser assisted pulp anesthesia for mandibular molar teeth with acute irreversible pulpitis is currently progressing.

## Conclusions

Laser irradiation applied directly on irreversibly inflamed pulp tissue for control of intra-operative pain was effective, thereby negating the need for additional local anesthetics. Post-operative pain evaluation showed laser activation was safe and did not cause any adverse discomfort.

## Data Availability

Raw data available for persons with research interest after a reasonable request

https://clinicaltrials.gov/ct2/show/NCT04735120?cntry=IN&city=Madurai&draw=2&rank=2

## Author Contributions

Conceptualization, Anand Sherwood; Formal analysis, Anand Sherwood; Investigation, Subashri V and Nivetha V; Methodology, Subashri V, Anand Sherwood, Paul Abbott and James Gutmann; Project administration, Anand Sherwood; Supervision, Omar Farooq and Aarthy MV; Writing – original draft, Anand Sherwood; Writing – review & editing, Paul Abbott and James Gutmann.

## Compliance with Ethical Standards

### Funding

The work received no external/internal funding of any nature.

### Ethical approval

All procedures performed in studies involving human participants were in accordance with the ethical standards of the institutional and/or national research committee and with the 1964 Helsinki declaration and its later amendments or comparable ethical standards.”

### Informed consent

Informed consent was obtained from all individual participants included in the study.

## Acknowledgement

The authors deny any conflicts of interest

## References

1. Rosenberg PA. Clinical strategies for managing endodontic pain. Endod Topics 2002; 3: 78–92.

2. Abbott PV, Parirokh M. Strategies for managing pain during endodontic treatment. Aust Endod J 2018; 44: 99–113.

3. Nivedha V, Sherwood AI, Abbott PV, et al. Pre-operative ketorolac efficacy with different anesthetics, irrigants during single visit root canal treatment of mandibular molars with acute irreversible pulpitis. Aust Endod J 2020; 46: 343–350.

4. Nagendrababu V, Pulikkotil SJ, Veettil SK, et al. Effect of nonsteroidal anti-Inflammatory drug as an oral premedication on the anesthetic success of inferior alveolar nerve block in treatment of irreversible pulpitis: A systematic review with meta-analysis and trial sequential analysis. J Endod 2018; 44: 914–922.

5. Nivedha V, Sherwood AI, Abbott PV, et al. The effect of pre-operative ketorolac tromethamine administration during single visit root canal treatment of mandibular molars with acute irreversible pulpitis on intra-/post-operative pain. Sci Arch Dent Sci 2021; 4(3): 3–14.

6. Chan A, Armati P, Moorthy AP. Pulsed Nd: YAG laser induces pulpal analgesia: a randomized clinical trial. J Dent Res 2012; 91(7Suppl): 79S–84S.

7. Ghabraei S, Chiniforush N, Bolhari B, et al. The effect of photobiomodulation on the depth of anesthesia during endodontic treatment of teeth with symptomatic irreversible pulpitis (Double blind randomized clinical Trial). J Lasers Med Sci 2018; 9: 11–14.

8. Abbott PV. Examination and diagnosis of the pulp, root canal periapical/periradicular conditions. In: Rotstein I, Ingle JI(eds) Ingle’s Endodontics, 7 ed. PMPHUSA, 2019; NC, US. pp. 215–225.

9. Nagendrababu V, Duncan HF, Bjørndal L, et al. PRIRATE 2020 guidelines for reporting randomized trials in Endodontics: explanation and elaboration. Int Endod J 2020; 53: 774–803

10. Bamini L, Sherwood A, Arias A, et al. Influence of tooth factors and procedural errors on the incidence and severity of post-endodontic pain: A prospective clinical study. Dent J (Basel) 2020; 8: 73.

11. Evangelin J, Sherwood IA, Abbott PV, et al. Influence of different irrigants on substance P and IL-8 expression for single visit root canal treatment in acute irreversible pulpitis. Aust Endod J 2020; 46: 17–25.

12. Goharkhay K, Moritz A, Wilder-Smith P, et al. Effects on oral soft tissue produced by a diode laser in vitro. LasersSurgMed 1999; 25:401–406.

13. Balasubramanian SK, Natanasabapathy V, Vinayachandran D. Clinical considerations of intrapulpal anesthesia in pediatric dentistry. Anesth Essays Res 2017;11: 1–2.

14. Ramalho KM, de Souza LM, Tortamano IP et al. A randomized placebo-blind study of the effect of low power laser on pain caused by irreversible pulpitis. LasersMed Sci 2016; 31: 1899–1905.

15. Naseri M, Asnaashari M, Moghaddas E, Vatankhah MR. Effect of low-level laser therapy with different locations of irradiation on postoperative endodontic pain in patients with symptomatic irreversible pulpitis: A double-blind randomized controlled trial. JLasers Med Sci 2020; 11: 249–254.

16. Arias A, de la Macorra JC, Hidalgo JJ, Azabal M. Predictive models of pain following root canal treatment: a prospective clinical study. Int Endod J 2013; 46: 784–793.

17. Strakas D, Franzen R, Kallis A. et al.. A comparative study of temperature elevation on human teeth root surfaces during Nd:YAG laser irradiation in root canals. Lasers Med Sci 2013; 28: 1441–1444.

